# Health Technology Assessment (HTA) readiness in Uganda: Stakeholder’s perceptions on the potential application of HTA to support National Universal Health Coverage efforts

**DOI:** 10.1101/2023.05.16.23290024

**Authors:** Chrispus Mayora, Joseph Kazibwe, Richard Ssempala, Brenda Nakimuli, Aloysius Ssenyonjo, Elizabeth Ekirapa, Sarah Byakika, Tom Aliti, Timothy Musila, Mohamed Gad, Anna Vassall, Francis Ruiz, Freddie Ssengooba

## Abstract

**Introduction:** Health technology assessment (HTA) is an area that remains less implemented in low- and lower middle-income countries. The aim of the study is to understand the perceptions of stakeholders in Uganda towards HTA and its role in decision making, in order to inform its potential implementation in the country.

**Methods:** The study takes a cross-sectional mixed methods approach, utilising an adapted version of an International Decision Support Initiative questionnaire with both semi-structured and open-ended questions. We interviewed thirty key informants from different stakeholder institutions in Uganda that have decision making roles in the health sector.

**Results:** All participants perceived HTA as an important tool for decision making. Allocative efficiency was regarded as the most important use of HTA receiving the highest average score (8.8 out of 10), followed by quality of healthcare (7.8/10), transparency (7.6/10), budget control (7.5/10) and equity (6.5/10). There was concern that some of the uses of HTA may not be achieved in reality if there was political interference during the HTA process. The technology areas that interviewees highlighted as needing HTA type evaluations urgently were identified as medicines (60.0% of the participants), diagnostics (53.3%), vaccines (40.0%), and public health programs (26.7%). The study participants identified development partners as the most likely potential users of HTA (66.7% of participants), followed by Ministry of Health (43.3%).

**Conclusion:** Interviewed stakeholders in Uganda viewed the role of HTA positively, suggesting that there exists a promising environment for the establishment and operationalisation of HTA as a tool for decision making within the health sector. However, sustainable development and application of HTA in Uganda will require adequate capacity both to undertake HTAs and to support their use and uptake.

## Introduction

Priority setting is a key aspect of attaining universal health coverage (UHC)^1^. UHC is commonly interpreted as people and communities receiving the health services they need without experiencing financial hardship no matter who they are or where they are^2^. Many low- and middle-income countries (LMICs) are aiming to achieve UHC by 2030, introducing reforms, for example, to remove financial barriers to care and eradicate the financial burden to the patients and their households. Some of the countries enacting UHC-relevant reforms include among others, Vietnam^3^, China^4^, Ghana^5^, Uganda^6^ and Zambia^7^.

Many UHC reforms in LMICs are focused at increasing the available resources for the health system while reducing out of pocket expenditure on health^8^. This leaves out the element of how the resources are spent, the purchasing function of the health financing building block of the health system. This includes the process by which funding priorities are made (priority setting for health). As countries grapple with increasing resource constraints with many emphasizing the need to maximise value for money, current institutional structures are often inadequate to support the efficient allocation and use of available resources^9^. Most decisions (including priority setting decisions) are made in ad hoc and implicit manner^10,11^. However, some countries have made progress in this space through incorporating evidence-informed decision-making (EIDM) approaches in health policies as a means to effectively and efficiently respond to the health needs of served populations. The World Health Organisation defines EIDM as a systematic and transparent approach that applies structured and replicable methods to identify, appraise and make use of evidence across decision-making processes, including for implementation^12^.

The WHO asserts that priority setting decisions should be informed by the best available evidence from research, as well as other factors such as context, public opinion, equity, feasibility of implementation, affordability, sustainability, and acceptability to stakeholders^12^. Countries in the global south have increasingly introduced EIDM mechanisms such as health technology assessment (HTA); or at least expressed a desire to do so^13–15^.

HTA has been defined as a multidisciplinary process that uses explicit methods to determine the value of a health technology at different points in its lifecycle^16^. The purpose is to inform decision-making in order to promote an equitable, efficient, and high-quality health system^17^. HTA has been highlighted by the WHO as one of the tools that can propel countries towards UHC if implemented. Examples of countries recently introducing HTA in decision making include Ghana^15,18^, India^19^, Kenya^20^, Indonesia^21^, and the Philippines^22^. Uganda is committed to the achievement of UHC by 2030 through strengthening the health system and its support mechanisms with a specific focus on primary health care^23^. The Ministry of Health (MoH) sees HTA as one of the tools that can enable the country make further progress towards UHC.

The International Decision Support Initiative (iDSI), a global network of priority setting support institutions and experts, received a request from the Ugandan MoH to facilitate the institutionalisation of HTA in the country^24^. Institutionalisation of HTA involves the establishment and operationalisation of HTA structures and processes that enable the sustainable production and utilisation of HTA evidence.

As an initial step, and to inform any strategy to implement HTA in Uganda, a situational (or landscape) analysis that examines existing priority setting approaches and the potential capacity to use evidence in decision making was considered necessary. The present study, a component of a wider situational analysis, aimed to understand the perceptions of stakeholders in Uganda towards HTA to inform HTA implementation in the country.

## Methods

### Study Design

This is a cross-sectional mixed methods study utilising a questionnaire with both semi-structured and open-ended questions.

### Context

The study was carried out in Uganda, a country located in East Africa. Uganda is a low income country with a gross domestic product per capita of USD 883.9^25^ and a population of 47 million as of 2021^26^. The Ugandan health system is decentralized from MoH to the districts but with the financing still largely controlled by the central government, there is little allocative authority available to the districts. The MoH is responsible for policy formulation, planning, quality assurance, epidemic response, international relations, resource mobilization and monitoring and evaluation. Most decision making takes place at the Ministry level. The budgetary allocations to the health sector have been increasing over time from 560 billion Uganda Shillings (UGX) in the financial year (FY) 2010/11 to 911 billion UGX in 2018/19^27^. However, the MoH budget as a share of the total government budget has been showing a downward trend even before COVID-19 pandemic. The budget declined from 11.2% in 2004/5 to 5.1% in 2020/21^28^. Out-of-pocket expenditure on health as a proportion of the national current health expenditure remains high at 40%^29,30^.

Uganda has a total of 6,937 health facilities and clinics at different levels in 128 districts^31^. The biggest share of all health facilities is government owned at 45.16% (3,133); another 14.44% (1,002) are Private and Not-For-Profit (PNFP), 40.29% (2,795) are Private for Profit (PFP) and 0.10% (7) community-owned facilities. Uganda has made improvements in healthcare coverage in recent years. The Government of Uganda planned to have at least 85% of the population living within five kilometres of a health facility by 2020 from 75% as of 2015^32^.This target however has not yet been achieved^33^.

### Data collection tool

The iDSI HTA situational analysis survey questionnaire^34^ was used to collect data from key informants. The questionnaire has previously been used in a survey of Sub-Saharan African countries and also in a more in depth study focused on Nigeria^35^. The questionnaire was adapted to the Uganda setting and used to collect the primary data. The adaptation of the tool was done during a one-day virtual workshop organised by the research team consisting of members from Makerere University School of Public Health (MakSPH), MoH and iDSI (co-authors on this study). The questionnaire is divided into two parts: a closed ended (quantitative) series of questions focused on the uses and importance of HTA; and an open-ended questionnaire section that seeks information on the need, demand and supply of HTA. The quantitative sections score the importance of HTA in a number of dimensions: achieving allocative efficiency; quality of care; transparency in decision making; budget control; and equity. The scores ranged from 0 to 10, where 0 is ‘not important’ and 10 is ‘very important’. The key informants were expected to answer both the quantitative and qualitative sections of the questionnaire. The questionnaire was pretested among 5 non-key informants at the MoH to ensure that the questions were clear and uniformly understood by the people answering them.

### Study participants

Participants were people in decision making positions within their organisations or institutions. They were purposively selected to represent a variety of stakeholder institutions involved in policy making in the health sector which are likely to utilise and supply HTA evidence. The selection of the key informants was based on expertise, experience, and role in the decision-making and resource-allocation/prioritization processes in the health sector at both regional and national level. The key informants were identified through a rapid literature review, and consultations with the MoH. The key informants included: representatives of the professional councils, for example the Uganda Medical and Dental Practitioners Council; District health officers; development partners; government institutions such as the National Drug Authority; investigators at academic and research institutions; and non-governmental organisations. The interviews took place between January and March 2022.

### Administration of the questionnaire

Appointments were made with the participants prior to the interview day. The interviews were conducted either in person or virtually via Zoom taking into account the key informant’s preference and the existing COVID 19 guidelines in the country at that time. The questionnaire was interviewer-administered. All interviews were recorded. In addition, the interviewer took notes during the interview which were handed over to the principal investigator after the interview for referencing during analysis.

### Data processing and analysis

The recorded interviews were transcribed. The quantitative data were cleaned and descriptively analysed using Microsoft Excel. The data was summarised narratively in addition to using tables and graphs.

The qualitative data was analysed using the inductive thematic analysis method^36^ taking the constructionist thematic approach. Validation of results was done through a meeting with the stakeholders.

### Ethical considerations

This study was reviewed and approved by the MakSPH Higher Degrees Research and Ethics Committee (HDREC), approval number SPH-2021-151. The study was registered with the Uganda National Council of Science and Technology. Ethical approval was also sought from and granted by the London School of Hygiene and Tropical Medicine Ethics Committee (LSHTM Ethics Ref: 26615). During data collection, ethical principles were upheld in the study including: 1) maintaining the level of confidentiality; 2) Informant’s participation was voluntary 3) obtaining written informed consent; and 4) no study materials contained names or other explicit identifiers of participants.

## Results

A total of thirty stakeholders from governmental, non-governmental and private sector institutions were interviewed to understand their perceptions towards the use of HTA in decision making. Supplementary material 1 shows the list of institutions from which the key informants where selected.

Proportionately, the key informants included in this study reflected a balanced representation of the supply and demand side of HTA in the country. The supply side of HTA was mostly represented by academic institutions (n=5) while the majority of key informants on the demand side were government ministries (n=8).

### Perceived use of HTA

The stakeholders perceived achieving allocative efficiency, quality of care, transparency in decision making, budget control and equity as key uses of HTA. Each of these aspects had average scores greater than six out of 10 (where 10 indicates the use is “very important”). Allocative efficiency was scored as the most important use of HTA gaining an average score of 8.8. Below is figure 1, a graph showing the average scores awarded to each of the HTA benefits.

**Figure 1:**
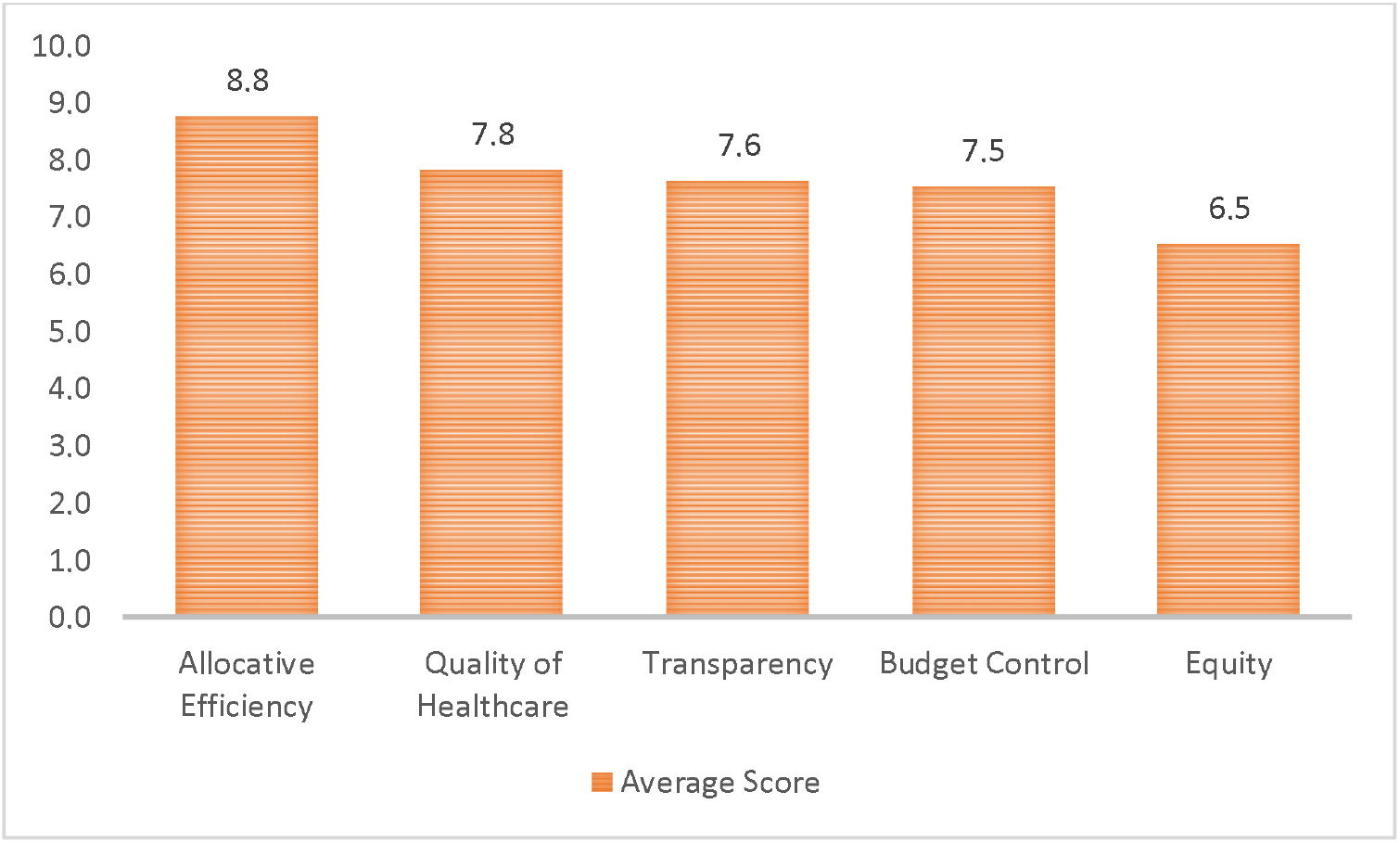
Perceived importance of HTA among stakeholders

Value for money and optimal use of resources were common reasons provided for scoring allocative efficiency highly. Health technology assessments were viewed by many as a way of getting the best value from the available resources enhancing the efficiency of the health system. *“Health Centre IV, we give resources in the same way that we give… other lower-level facilities, and yet you know very well that almost 30% of these health centres are not carrying out operations but they are getting money like the health centre IVs which are having like 60 or 50 operations per month. So, there you are not allocating your resources efficiently. There is no allocative efficiency in that aspect. So, it means that you need to have that information and you need to develop that criterion, the whole of that formula then you can be able to say that you can efficiently allocate, we must be conscious of what we are doing….”* key informant (K3)

Others emphasized the need for transparency in decision making to ensure that the resourc allocation is aligned with the main problems from the communities’ perspective. A key reason for adopting HTA was that it would help improve the quality of health care.

*“Improving the quality of healthcare, ……. requires having in place tools that can help you to continuously assess and identify the gap on a regular basis. So, if you go manual, you cannot get all the information and analyse it in time and triangulate and give you a good assessment of the different perspectives of quality…. HTA can assist in ensuring provision of quality services”* Key informant (K9)

*“why would I give it [improving quality of healthcare] a higher score… because interventions implemented based on evidence are less prone to embezzlement. …, resources are put to their best use.”* Key informant (K23).

Despite a high score on the *need* for HTA, many respondents acknowledged that there could be a less-than-optimal use of HTA in actual decision making due to political pressures and other considerations. For example, limited resources and capacity may not support the actual use of evidence in practice.

*“I will give it about 8 (allocative efficiency), because as much as you can have the assessment [HTA] done,…when it comes to decision making, there are other factors that also influence things like political decisions, interests and all that. It is good, but you cannot rely on it 100%”.* Key informant (K9).

### Areas that require HTA urgently

Each respondent was asked to identify three technology (or intervention) areas that they thought urgently required HTA. A number of areas were identified including medicines, medical devices and diagnostics, public health programs, vaccines, screening programs, and service delivery incentives (Figure 2).

**Figure 2:**
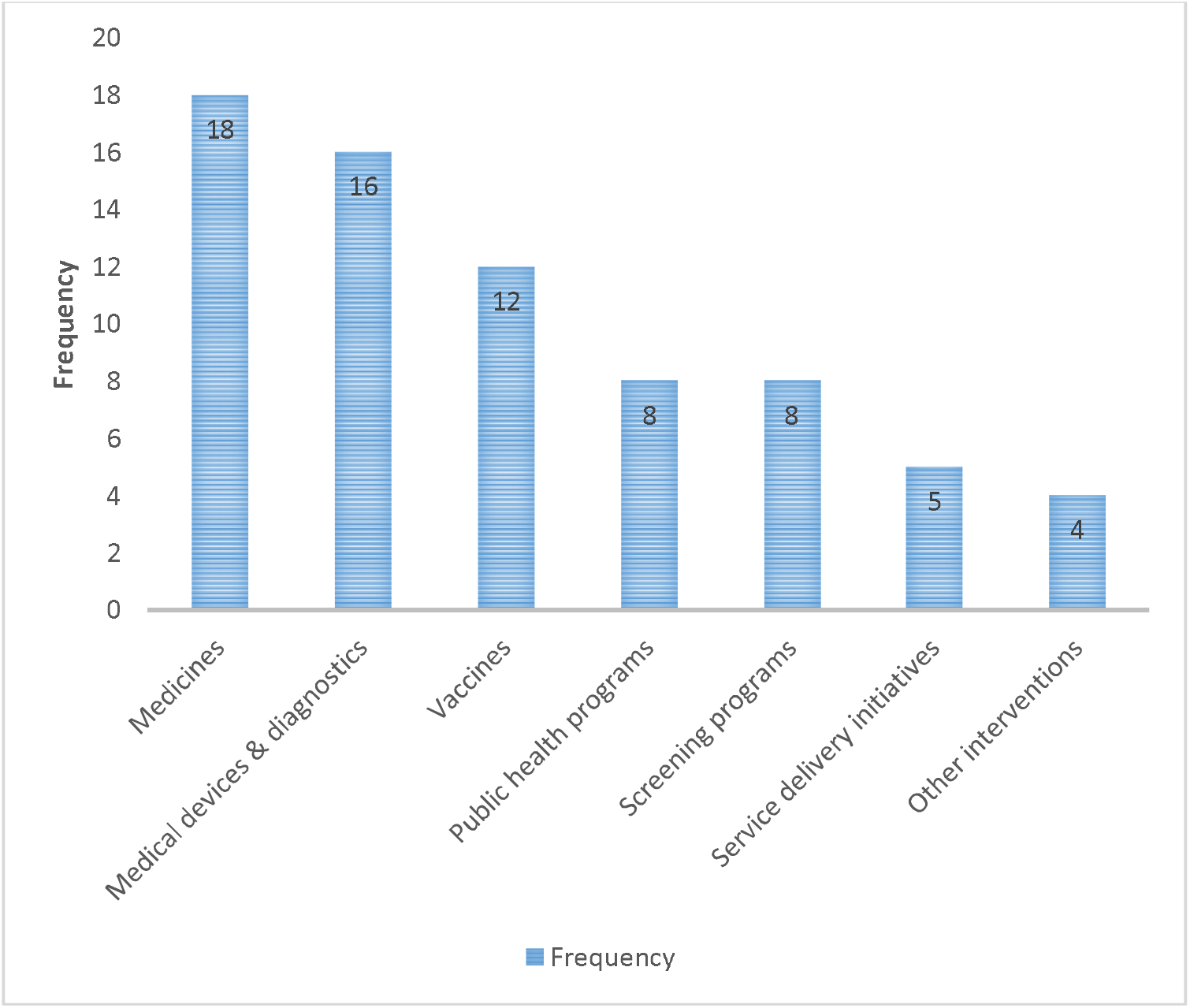
Areas that require the use of HTA urgently

The assessment of medicines was identified as the area HTA was most urgently needed (18 respondents [60.0%]), followed by medical devices and diagnostics (16 respondents [53.3%]), and vaccines (12 respondents [40.0%]) in third place (figure 2).

Most respondents highlighted that these technology areas are expensive relative to the limited available resources. Other issues noted by respondents included the use of expensive first line drugs by Ugandan health providers, especially in the private sector, despite the availability of more affordable alternatives.

*“…. these are things (Medicines, vaccines, and medical devices) which are common in a health system service delivery and because they are there, we interface with them whether you are a patient or not. […] The diagnostic devices [….], people are taken advantage of with these devices so I think to help the user, these need to go through HTA process and government or the country makes a statement on the various diagnostics; then the private sector is unlikely to take advantage of the population ……”* Key informant (K10)

### Potential users of HTA outputs

Respondents were asked for the likely users of HTA outputs. Figure 3 shows the frequency of perceived likely users of HTA outputs in Uganda.

**Figure 3:**
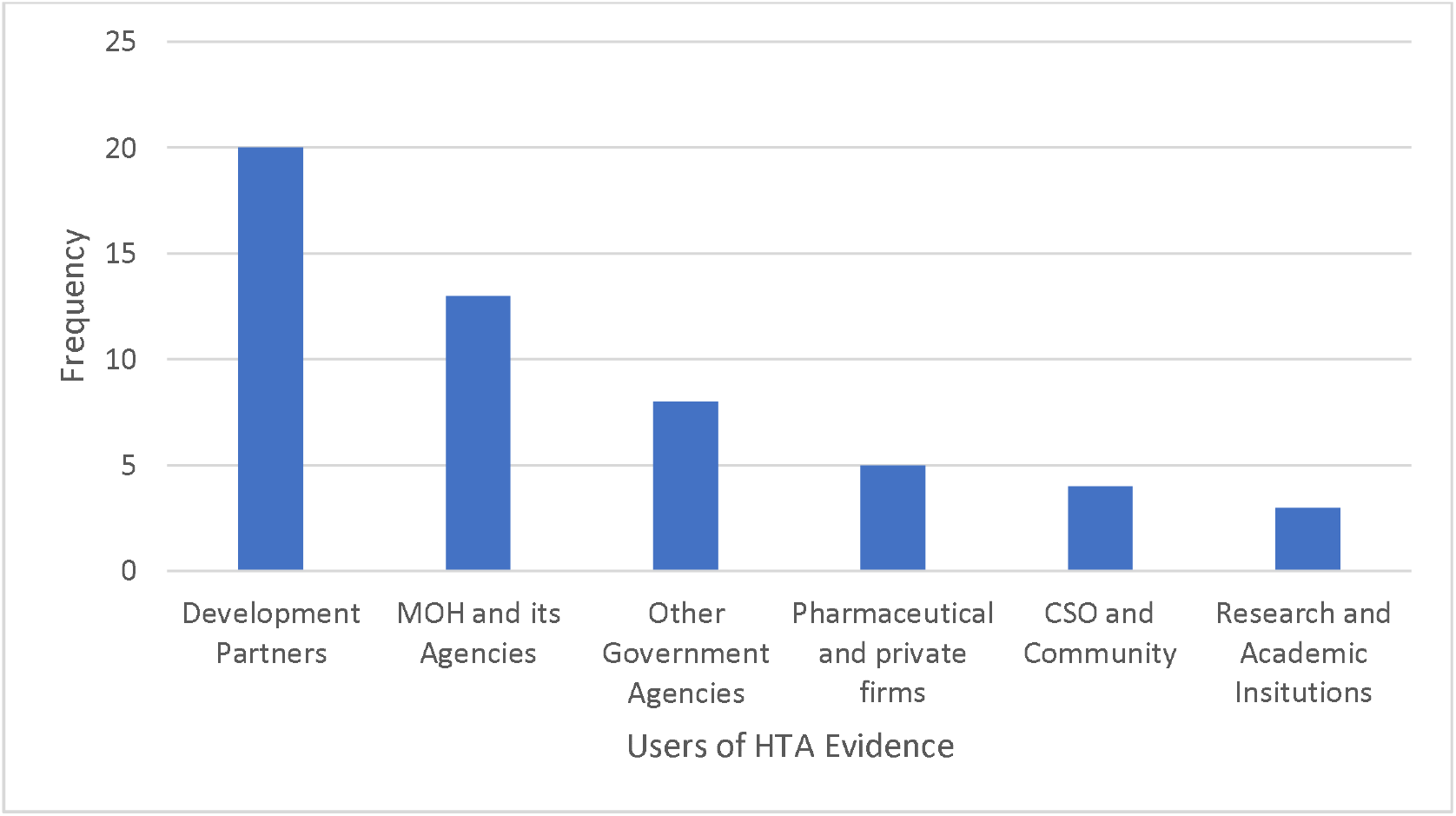
Likely users of HTA evidence

Development partners were perceived to be most likely users of HTA output (20 of 30 respondents) followed by the MoH and its subsidiaries (13 of 30). Only five respondents perceived pharmaceutical and private firms as potential users of HTA outputs, while civil society organisations (CSO), community and research institutions were seen as potential users of HTA output by less than five respondents.

### Perceived level and type of HTA evidence needed by major stakeholders

Respondents scored the extent to which the different type of HTA outputs are needed by the major categories of stakeholders (figure 4).

**Figure 4:**
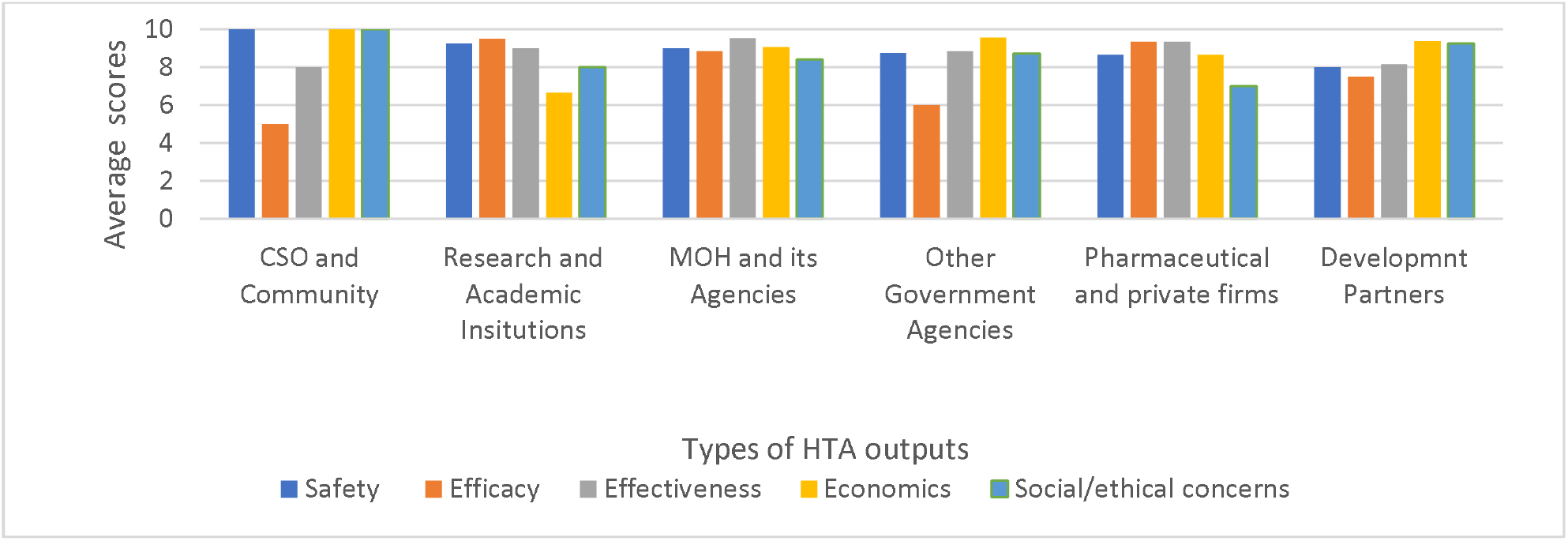
Perceived need for the different types of HTA outputs by stakeholders

All the different potential HTA outputs (relating to safety, efficacy, effectiveness, economic and social/ethical concerns) are seen to be needed and helpful to all categories of stakeholders (table 1). All the outputs were scored at least five out of ten for each category of stakeholders. ‘Efficacy’ was scored at 5/10 for the CSO and community stakeholders.

**Table 1:** Level of interest in different types of HTA outputs

| Organization | Safety | Efficacy | Effectiveness | Economics | Social/Ethical |
| --- | --- | --- | --- | --- | --- |

|  |  |  |  |  | **concerns** |
| --- | --- | --- | --- | --- | --- |
| Development Partners | 8.00 | 7.50 | 8.14 | 9.37 | 9.25 |
| MOH and its Agencies (NMS, UNITAG, NDA) | 9.00 | 8.85 | 9.53 | 9.06 | 8.41 |
| Other Government Agencies (MoFPED, NPA, KCCA, UBOS) | 8.75 | 6.00 | 8.83 | 9.57 | 8.71 |
| Pharmaceutical and private firms (JMS) | 8.67 | 9.33 | 9.33 | 8.67 | 7.00 |
| CSO and Community | 10.00 | 5.00 | 8.00 | 10.00 | 10.00 |
| Research and Academic Institutions | 9.25 | 9.50 | 9.00 | 6.67 | 8.00 |

Source: Averaged from respondent scores

The CSO category of stakeholders was perceived to be the most interested in safety related outputs of HTA (10.00/10) followed by research and academic institutions (9.25/10), and the MoH and its subsidiaries (9.00/10). Research and academic institutions were identified as the most interested in efficacy aspects (9.50/10) followed by pharmaceutical and private firms (9.33/10). The MoH and its subsidiaries was identified as the stakeholder most interested in effectiveness outputs of HTA (9.53/10) followed by pharmaceutical and private firms (9.33/10). The CSO were perceived to be the most interested in economic-related HTA outputs (10 out of 10), followed by development partners (9.37/10). The ranking of the interest of the stakeholders in social/ethical concerns was similar to the interest in economic evidence.

## Discussion

This study shows that HTA stakeholders (decision makers) on both the supply and demand sides of HTA perceive HTA as an important tool for decision making within the health sector in Uganda. According to this study, seeking allocative efficiency is the most important goal when implementing HTA, and medicines were identified as the main technology area where the application of HTA type analyses were long overdue. Notably, development partners were perceived to be the most likely users of HTA outputs.

Resource allocation was perceived as the most prominent use of HTA according to the stakeholders in Uganda. This is consistent with findings in India^34^ and Nigeria^35^ where similar surveys have been done. HTA has been taken to be synonymous with cost effectiveness^37^ and limited to the end result of resource allocation or some assessment of value-for-money. However, HTA arguably offers further benefits in terms of transparency, support for equity considerations and stakeholder inclusiveness, as part of an overarching priority setting decision process^38–40^. HTA is not a narrow technical exercise; it provides a framework to accommodate multiple considerations/criteria including potentially ‘political factors’ within a multi-stakeholder engagement process. A good HTA process follows pre-agreed rules and offers transparency with respect to how any evidence is considered in decision making. These factors enhance the credibility and social legitimacy of often difficult priority setting choices^41^.

Development partners were identified as the most likely users of HTA in Uganda in this study which differs from the findings of similar studies in other countries. In Nigeria and India, it was found that that federal and state governments were seen as the dominant users of HTA outputs^34,35^. This difference could reflect variation in development partner influence in resource allocation decisions in these settings. In both Nigeria and India, the state governments have extensive authority to make such decisions, while in Uganda, most of the health care resources have been ear-marked already, and to a significant extent these ear-marked resources are supported by funding from development partners. Indeed, development partners fund approximately 40% of the Ugandan health budget^30^. The findings from Uganda may also reflect the stakeholders’ association of HTA with primarily the allocation of resources for specific technologies, with less consideration of other uses such as informing the development of clinical guidelines. Further sensitisation of Ugandan stakeholders on the nature and use of HTA may change perceptions around key beneficiaries of implementation.

It is perhaps concerning that the MoH is not seen as the most likely user of HTA evidence despite the fact that it is the institution responsible for policy formulation and implementation within the health sector. This may in part be a result of the relatively low engagement of stakeholders by the MoH on the topic of HTA. This suggests that there could be value in the MoH (with the support of international partners as needed) in actively engaging with relevant stakeholders, and advocate for a potential legal framework to guide the operationalisation and institutionalisation of HTA in the country. Building HTA within a legal framework that requires it use for coverage decisions could be valuable in aiding implementation, especially in environments where there is a national social insurer^42^. For example, the Philippines put in place an HTA organisational structure informed by a statutory law that established the Health Technology Assessment Council (HTAC), an independent advisory institution which gives guidance to the Philippines Government Department of Health and the Philippine Health Insurance Corporation (PhilHealth) on which health interventions/technologies are to be funded by the government^22^.

Uganda, through the MoH, is in fact pursuing a legal framework detailing the processes through which priority setting decisions are made and operationalised. However significant progress can still be made in the absence of a detailed legal underpinning for HTA. For example, Thailand has developed robust HTA systems without a specific legal framework and has created a semi-autonomous unit to serve as an agency to inform decisions using HTA^43^. Rwanda, while not having a dedicated HTA unit, is currently applying HTA approaches to update the health benefits package of the community-based health insurance scheme^44^. Uganda could therefore explore setting up preliminary, exploratory structures to support HTA development, such as an HTA unit perhaps within the MoH, in advance of a formal legal framework. This would allow for testing potential options and processes.

The relative absence of HTA-like approaches in decision making in Uganda was seen by the stakeholders interviewed as disadvantaging the patients and people in Uganda in general. The cost of medicines is not regulated and patients are seen as being taken advantage of. This potentially exposes patients and their households to catastrophic health expenditures. It is for these reasons that respondents see HTA as a tool to support development of the national drug formulary, standardize reimbursement of expenses especially for provider payment systems and also improve health outcomes of the final users.

The establishment of HTA structures is likely to stimulate and increase HTA capacity in the country. Currently, there is a limited HTA relevant literature that focuses on the Ugandan context, and the majority of those studies are authored by researchers based in other countries, a situation similar to LMICs more generally^45^. The existence of HTA structures will further enhance the value of the awaited Master of Health Economics program by Makerere University which will train health economists in-country that could then be absorbed into those HTA structures.

Building in-country HTA structures will however still need the support of international organisations and donors for the foreseeable future. There are a number of development partners that are engaging in HTA relevant activities within the country including the Medical Research Council/Uganda Virus Research Institute and LSHTM Uganda Research Unit, the University of York under Thanzi la Onse, the Professional Society for Health Economics and Outcomes Research (ISPOR), Results for Development (R4D), ThinkWell, Strategic Purchasing Africa Resource Centre (SPARC), Norwegian Institute of Public Health (NiPH), and KEMRI-Wellcome Trust. Although important for supporting domestic HTA institutionalisation, in most cases development partners tend to carry out activities in isolation leading to duplication and unnecessary competition if unregulated within a given country. There is therefore a need for the MoH to encourage collaboration and coordination among these stakeholders to avoid duplication of HTA related activities and optimize impact.

## Limitations

The study included respondents that were in decision making positions within their organizations. The findings may not reflect mid-level and low-level managers within those institutions. The stakeholders selected were mostly from the central region of Uganda which is generally urban and houses the headquarters of MoH and most key institutions. Therefore, the findings of this study may not reflect the perceptions of decision makers that are based in rural parts of the country. The level of understanding of HTA varied across stakeholders, where those with a background in health economics having more knowledge on the subject matter than others. This may have introduced some level of bias in the findings.

## Conclusion

Key stakeholders in Uganda took a positive view of the role of HTA suggesting a promising environment for the establishment and operationalisation of HTA as a tool for decision making within the health sector. Sustainable development and application of HTA in Uganda will require adequate capacity both to undertake HTAs and to support their use and uptake. There is perhaps a need for a more comprehensive understanding of current HTA capacity in Uganda, which takes into account the different needs and requirements of the stakeholders involved, the existing priority setting processes and notes the importance of strengthening both technical and non-technical (e.g. administrative) aspects necessary for the conduct of HTA. Supported by development partners and under the leadership of Ugandan authorities, it may be necessary to undertake a detailed capacity assessment exercise in order to better understand existing strengths and where current expertise is located. This would inform a national strategy for capacity building in HTA going forward.

## Data Availability

All data produced in the present study are available upon reasonable request to the authors

## Declaration

### Ethics approval and consent to participate

This study was reviewed and approved by the MakSPH Higher Degrees Research and Ethics Committee (HDREC), approval number SPH-2021-151 and the London School of Hygiene and Tropical Medicine Ethics Committee (LSHTM Ethics Ref: 26615).

## Consent for publication

Not applicable

## Availability of data and materials

The data analysed during the current study is available from the corresponding author upon reasonable request.

## Authors contributions

Conceptualisation: – MG, JK, FR, FS, CM

Developing protocol: – JK, CM, RS, AS, EE, SB, TA, TM, AV, FR

Data collection: – RS, BN

Validation: – JK, FR, EE

Data curation and analysis: – JK, RS, BN

Funding acquisition: – FR, AV

Methodology: – JK, CM, FR, AV

Project administration: – JK, MG

Supervision: – FR, FS

Writing – original draft: – JK, CM, RS

writing review & editing: – JK, CM, RS, BN, AS, EE, SB, TA, TM, MG, AV, FR, FS

## Acknowledgements

None

## References

1. WHO Consultative Group on Equity and Universal Health Coverage. Making Fair Choices on the Path to Universal Health Coverage.; 2014. Accessed September 2, 2022. http://www.ncbi.nlm.nih.gov/pubmed/25666865

2. World Health Organisation. Universal health coverage (UHC). Published April 1, 2021. Accessed June 12, 2022. https://www.who.int/news-room/fact-sheets/detail/universal-health-coverage-(uhc)

3. Tran Van Tien by, Thi Phuong H, Mathauer I, Thi Kim Phuong N. A Health Financing Review of Vietnam with a Focus on Social Health Insurance: Bottlenecks in Institutional Design and Organizational Practice of Health Financing and Options to Accelerate Progress towards Universal Coverage.; 2011.

4. Zhang J, Zeng H, Yu T, Zhang R. China’s universal healthcare reform: the first phase [2009–2011] of the ambitious plan. J Hosp Manag Heal Policy. 2018;2:22–22. doi:10.21037/JHMHP.2018.04.09

5. Fusheini A. Healthcare Financing Reforms: Ghana’s National Health Insurance. Heal Reforms Across World. Published online March 2020:25–54. doi:10.1142/9789811208928_0002

6. Cashin C, Dossou J-P. Can National Health Insurance Pave the Way to Universal Health Coverage in Sub-Saharan Africa? https://doi.org/101080/2328860420212006122. 2021;7(1). doi:10.1080/23288604.2021.2006122

7. Masiye F, Chansa C. Health Financing in Zambia.; 2019.

8. Jalali FS, Bikineh P, Delavari S. Strategies for reducing out of pocket payments in the health system: a scoping review. Cost Eff Resour Alloc. 2021;19(1):1–22. doi:10.1186/S12962-021-00301-8/TABLES/5

9. Glassman A, Chalkidou K, Giedion U, et al. Priority-Setting Institutions in Health: Recommendations from a Center for Global Development Working Group. Glob Heart. 2012;7(1):13–34. doi:10.1016/J.GHEART.2012.01.007

10. Baltussen R, Niessen L. Priority setting of health interventions: The need for multi-criteria decision analysis. Cost Eff Resour Alloc. 2006;4(1):1–9. doi:10.1186/1478-7547-4-14/FIGURES/2

11. Chalkidou K, Levine R, Dillon A. Helping poorer countries make locally informed health decisions. BMJ. 2010;341(7767):284-286. doi:10.1136/BMJ.C3651

12. World Health Organsiation. Evidence, Policy, Impact: WHO Guide for Evidence-Informed Decision-Making.; 2022. Accessed December 14, 2022. https://www.who.int/publications/i/item/9789240039872

13. Downey LE, Mehndiratta A, Grover A, et al. Institutionalising health technology assessment: establishing the Medical Technology Assessment Board in India. BMJ Glob Heal. 2017;2(2):e000259. doi:10.1136/BMJGH-2016-000259

14. MacQuilkan K, Baker P, Downey L, et al. Strengthening health technology assessment systems in the global south: a comparative analysis of the HTA journeys of China, India and South Africa. Glob Health Action. 2018;11(1). doi:10.1080/16549716.2018.1527556/SUPPL_FILE/ZGHA_A_1527556_SM6628.ZIP

15. Groom G, Groom, Genevive. Final Report: iDSI learning review: Ghana. F1000Research 2019 8840. 2019;8:840. doi:10.7490/F1000RESEARCH.1116868.1

16. World Health Organisation. International HTA networks. Accessed April 15, 2021. https://www.who.int/health-technology-assessment/networks/en/

17. O’Rourke B, Oortwijn W, Schuller T. The new definition of health technology assessment: A milestone in international collaboration. Int J Technol Assess Health Care. 2020;36(3):187–190. doi:10.1017/S0266462320000215

18. Bidonde J, Meneses-Echavez JF, Asare B, et al. Developing a tool to assess the skills to perform a health technology assessment. BMC Med Res Methodol. 2022;22(1):78. doi:10.1186/S12874-022-01562-4/FIGURES/3

19. Ministry of Health and Family Welfare Department of Health Research India. Health Technology Assessment in India (HTAIn). Published 2018. Accessed April 30, 2023. https://htain.icmr.org.in/

20. Ministry of Health. Government launches health technology assessment to inform policy decision making Nairobi, Kenya. Published March 2018. Accessed April 30, 2023. https://www.health.go.ke/government-launches-health-technology-assessment-to-inform-policy-decision-making-nairobi-kenya-18-march-2018/

21. Sharma M, Teerawattananon Y, Luz A, Li R, Rattanavipapong W, Dabak S. Institutionalizing Evidence-Informed Priority Setting for Universal Health Coverage: Lessons From Indonesia. Inq (United States). 2020;57. doi:10.1177/0046958020924920

22. Republic of Philippines Department of Health. Health Technology Assessment Council (HTAC). Accessed April 30, 2023. https://hta.doh.gov.ph/health-technology-assessment-council-htac/

23. Ministry of Health. Ministry of Health Strategic Plan 2020/21 - 2024/25 - Ministry of Health | Government of Uganda. Published 2020. Accessed September 2, 2022. https://www.health.go.ug/cause/ministry-of-health-strategic-plan-2020-21-2024-25/

24. International Decision Support Initiative (iDSI). iDSI. Accessed September 6, 2022. https://www.idsihealth.org/

25. The World Bank. GDP per capita (current US$) - Uganda. Published 2022. Accessed April 30, 2023. https://data.worldbank.org/indicator/NY.GDP.PCAP.CD?locations=UG

26. The World Bank. Uganda. Published 2022. Accessed December 14, 2022. https://data.worldbank.org/country/UG

27. Ministry of Finance Planning and Economic Development. Good Governance; a Prerequisite to Harness the Demographic Dividend for Sustainable Development. State of Uganda Population Report 2018.; 2018.

28. UNICEF. The National Budget Framework Paper 2020/21 Budget Brief.

29. Kadowa I. A Case Study of the Uganda National Minimum Healthcare Package the Role of Essential Health Benefits in the Delivery of Integrated Services: Learning from Practice in East and Southern Africa.; 2017.

30. Ministry of Health Uganda. National Health Accounts 2016-2019.; 2022.

31. Ministry of Health of the Republic of Uganda. National Health Facility Master List.; 2018.

32. Ministry of Health Uganda. Health Sector Development Plan 2015/16 - 2019/20. Published 2015. Accessed January 8, 2022. https://www.health.go.ug/cause/health-sector-development-plan-2015-16-2019-20/

33. World Health Organisation African Region. Uganda on the right path to achieving Universal Health Coverage. Accessed April 30, 2023. https://www.afro.who.int/news/uganda-right-path-achieving-universal-health-coverage

34. Dabak SV, Pilasant S, Mehndiratta A, et al. Budgeting for a billion: Applying health technology assessment (HTA) for universal health coverage in India. Heal Res Policy Syst. 2018;16(1):1–7. doi:10.1186/S12961-018-0378-X/TABLES/2

35. Uzochukwu BSC, Okeke C, O’Brien N, Ruiz F, Sombie I, Hollingworth S. Health technology assessment and priority setting for universal health coverage: A qualitative study of stakeholders’ capacity, needs, policy areas of demand and perspectives in Nigeria. Global Health. 2020;16(1):1–11. doi:10.1186/S12992-020-00583-2/TABLES/6

36. Braun V, Clarke V. Using thematic analysis in psychology. Qual Res Psychol. 2006;3(2):77–101. Accessed January 26, 2023. https://uwe-repository.worktribe.com/preview/1043068/thematic_analysis_revised_-_final.pdf

37. Teerawattananon Y, Painter C, Dabak S, et al. Avoiding health technology assessment: a global survey of reasons for not using health technology assessment in decision making. Cost Eff Resour Alloc. 2021;19(1):1–8. doi:10.1186/S12962-021-00308-1/FIGURES/3

38. Pan American Health Organisation and World Health Organisation. Health Technology Assessment (HTA). Published 2013. Accessed April 30, 2023. https://www3.paho.org/hq/index.php?option=com_content&view=article&id=9229:2013-tecnologias-sanitarias&Itemid=41687&lang=en#gsc.tab=0

39. Gagnon MP, Desmartis M, Lepage-Savary D, et al. Introducing patients’ and the public’s perspectives to health technology assessment: A systematic review of international experiences. Int J Technol Assess Health Care. 2011;27(1):31–42. doi:10.1017/S0266462310001315

40. Gagnon MP, Tantchou Dipankui M, Poder TG, Payne-Gagnon J, Mbemba G, Beretta V. Patient and public involvement in health technology assessment: update of a systematic review of international experiences. Int J Technol Assess Health Care. 2021;37(1):e36. doi:10.1017/S0266462321000064

41. Rocchi A, Chabot I, Glennie J. Evolution of health technology assessment: best practices of the pan-Canadian Oncology Drug Review. Clin Outcomes Res. 2015;7:287–298. doi:10.2147/CEOR.S82549

42. Bertram M, Dhaene G, Edejer TT-T. Institutionalizing Health Technology Assessment Mechanisms: A How to Guide.; 2021. Accessed May 13, 2023. https://apps.who.int/iris/handle/10665/340722

43. Sharma M, Teerawattananon Y, Dabak SV, et al. A landscape analysis of health technology assessment capacity in the Association of South-East Asian Nations region. Heal Res Policy Syst. 2021;19(1):1–13. doi:10.1186/S12961-020-00647-0/FIGURES/3

44. Government of Rwanda. Rwanda Ministerial Instructions determining the methodology to define the community-based health insurance benefit package. Published August 31, 2021. Accessed April 27, 2022. https://gazettes.africa/archive/rw/2021/rw-government-gazette-dated-2021-08-31-no-special.pdf

45. Hollingworth S, Fenny AP, Yu SY, Ruiz F, Chalkidou K. Health technology assessment in sub-Saharan Africa: a descriptive analysis and narrative synthesis. Cost Eff Resour Alloc. 2021;19(1):1–13. doi:10.1186/S12962-021-00293-5/TABLES/2

