## Supplementary material 1 for "Health Technology Assessment (HTA) readiness in Uganda: Stakeholder’s perceptions on the potential application of HTA to support National Universal Health Coverage efforts"

Table 1: Affiliations of the key informants included in this study

| **Category** | **Institution** | **Number of key informants (N=30)** | **Percentage (%)** |
| --- | --- | --- | --- |
| **HTA supply side** | Academic institutions (both public and private universities) | 5 | 16.7 |
|  | Government health research departments/institutions | 1 | 3.3 |
|  | Professional Associations in health economics (International Health Economics Association Uganda Chapter) | 1 | 3.3 |
|  | Private research firms (e.g Consultancy firms) | 2 | 6.7 |
|  | Non-Governmental Organizations and Development partners | 4 | 13.3 |
| **Demand side for HTA** | Government ministries (eg MoH and Ministry of Finance) | 8 | 26.7 |
|  | Government departments responsible for procurement and regulation of medicines (e.g National Medical Stores and National Drug Authority) | 2 | 6.7 |
|  | Regional governments | 1 | 3.3 |
|  | Pharmaceutical companies | 1 | 3.3 |
|  | Professional Councils (Medical and Dental Practitioners Council and Allied Health Professionals Council) | 3 | 10.0 |
|  | NGOs and development partners | 2 | 6.7 |
